# Sagittal balance of the spine - lumbar lordosis or lumbosacral lordosis?

**DOI:** 10.1101/2024.03.19.24304424

**Authors:** Kai Song, Pengfei Chi, Qiang Yang, Cao Yang, Bing Wang, Fangcai Li, Zezhang Zhu, Weishi Li, Jianguo Zhang, Zheng Wang

**Affiliations:** Department of Orthopaedics, Chinese People’s Liberation Army General Hospital, Beijing, PR China; Department of Spine Surgery, Tianjin Hospital, Tianjin, PR China; Department of Orthopaedics, Union Hospital, Tongji Medical College, Huazhong University of Science and Technology, Wuhan, Hubei, PR China; Department of Spine Surgery, Second Xiangya Hospital of Central South University, Changsha, Hunan, PR China; Department of Orthopedics Surgery, Second Affiliated Hospital, School of Medicine, Zhejiang University, Hangzhou, Zhejiang, PR China; Department of Spine Surgery, The Affiliated Drum Tower Hospital of Nanjing University Medical School, Nanjing, Jiangsu, PR China; Department of Orthopaedics, Peking University Third Hospital, Beijing, PR China; Department of Orthopedics, Peking Union Medical College Hospital, Beijing, PR China; Shenzhen Institute of Advanced Technology, Chinese Academy of Sciences, Shenzhen, PR China

**Keywords:** Sagittal spinal alignment, Sagittal spinal curvature, Lumbosacral lordosis, Pelvic incidence, Maximal thoracolumbar vertebral tilt

## Abstract

**Objective:** To investigate sagittal spinal alignment from the perspective of the overall curvature of the “S” curve of the human spine, and explore the roles of pelvic incidence (PI) and maximal thoracolumbar vertebral tilt(TLmax) in the classification of the sagittal spinal aligment.

**Methods:** The tilt of the sacral, lumbar, and thoracic vertebrae (from Co1, S5, S4… to C7) were measured. The minimal sacral vertebral tilt(Smin), maximal thoracolumbar vertebral tilt(TLmax), and minimal thoracic vertebral tilt (Tmin) were recorded. The concept of lumbosacral lordosis (LSL) was introduced, and the Ferguson method was utilized to measure sagittal spinal parameters both in anatomical segmentation (Ferguson L1-S2, Ferguson T1-T12) and functional segmentation (Ferguson LSLmax, Ferguson TKmax). The subjects were grouped based on pelvic incidence (PI) and TLmax separately, and the mean and standard deviation of each parameter were calculated. Chi-square tests were conducted for statistical analysis.

**Results:** 1. Based on PI grouping: PI for all subjects was 45.4 ± 9.5° (21.7-86.4°). Group A consisted of 117 subjects with a mean PI of 34.7 ± 4.4°, Group B had 158 subjects with a mean PI of 45.2 ± 2.9°, and Group C included 113 subjects with a mean PI of 56.7 ± 5.8°. No statistically significant differences were found in tilt of S2, L1, T1, TLmax, Tmin, and Ferguson L1-S2, Ferguson T1-T12, and Ferguson TKmax among Groups A, B, and C. 2. Based on TLmax grouping: TLmax for all subjects was 110.5 ± 5.5° (94.4-132.0°). Group A had 91 subjects with a mean TLmax of 104.0 ± 2.3°, Group B comprised 216 subjects with a mean TLmax of 110.2 ± 2.1°, and Group C included 81 subjects with a mean TLmax of 118.6 ± 3.8°. Significant statistical differences were observed in tilt of S2, L1, T1, Smin, TLmax, Tmin, and Ferguson L1-S2, Ferguson T1-T12, Ferguson LSLmax, and Ferguson TKmax among Groups A, B, and C.

**Conclusion:** There were no differences in the magnitude of LSL and TK among subjects with different PI, indicating that PI does not affect the overall curvature of the “S” curve in the sagittal spinal aligment. In contrast, TLmax effectively distinguishes the overall curvature of the “S” curve.

## Introduction

The recognition of sagittal spinal alignments is of significant importance for the diagnosis and treatment of spinal disorders [1, 2]. The “S” curve formed by the lumbar lordosis and thoracic kyphosis has played a crucial role in the evolutionary process of humans transitioning from quadrupedal to bipedal upright locomotion [3-6]. Typically, scholars define the lumbar lordosis (LL) as the Cobb angle from L1 to S1 and the thoracic kyphosis (TK) as the Cobb angle from T1 to T12, to anatomically segment and analyze the “S” curve in radiographic images [7].

In recent years, there has been a preference among scholars to use functional segmentation based on the curvature of the “S” curve, rather than rigidly using the upper endplate of L1 as the boundary between the two bends. This approach is similar to the concept of the end vertebrae commonly used in scoliosis, where the inflection point’s position is not determined by the anatomical features of the vertebrae but by the characteristics of the spinal sequence [8-11]. Consequently, the cephalic end of LL may be located either proximally or distally on L1. In contrast, the caudal starting point of the lordosis is consistently defined by the upper endplate of S1.

Contrary to the previous study that considered the sacrum as a whole kyphotic segment [12-14], Song et al. confirmed that, in terms of spinal sequence, the sacrum is not entirely kyphotic but is divided into a lordotic segment (S1-S2) and a kyphotic segment (S2-S5, Co1). S1 and S2 actually continue the LL, and S2 should be considered the caudal vertebra of the lordotic segment in the sagittal spinal aligment [15]. If this is the case, the traditional definition of the “S” curve is incomplete, as it lacks the sacral lordotic segment at its distal end. In the context of studying spinal sequences, the definition of lumbosacral lordosis (LSL) seems more reasonable than lumbar lordosis alone.

Based on this background, the present study aims to measure and analyze the spinal curvature of the “S” curve in the general population from both anatomical segmentation and functional segmentation, using vertebral tiltand the Ferguson method. The study also considers the maximum thoracolumbar tilt (TLmax) as a reference for classification and typing of the sagittal spinal curvature, comparing it with previous classification types based on pelvic incidence (PI) [9-11, 16-19]. This comparison aims to provide a deeper exploration of the characteristics of the human sagittal spinal aligments and offer references for the diagnosis and treatment of spinal disorders.

## Methods

The study included 388 healthy adults aged 18-35 years old and collected their full-length lateral spine X-rays in a natural standing position. Among them, 336 cases were from Chinese PLA General Hospital, where they had been suspected of having scoliosis by other hospitals, but subsequent outpatient imaging examinations confirmed no abnormalities; 52 cases were from collaborating hospitals. Ethics committee of Chinese PLA General Hospital gave ethical approval (S2023-446-02) for this work.

### 1. Measurements

The tilt (right angle to the horizontal line) of the anterior, middle, and posterior edges of the vertebrae from Co, S5, S4… to C7 were measured. The minimal sacral vertebral tilt(Smin), the maximal thoracolumbar vertebral tilt(TLmax), and the minimal thoracic vertebral tilt(Tmin) were recorded. The lumbosacral lordosis angle (Ferguson L1-S2), thoracic kyphosis angle (Ferguson T1-T12), maximal lumbosacral lordosis angle (Ferguson LSLmax, calculated as Ferguson Smin-TLmax), and maximal thoracic kyphosis angle (Ferguson TKmax, calculated as Ferguson TLmax-Tmin) were calculated.

### 2. Conventional measurements

Pelvic incidence (PI), sacral slope (SS), pelvic tilt (PT), lumbar lordosis angle (LL, Cobb L1-S1), thoracic kyphosis angle (TK, Cobb T1-T12), T9 spino-pelvic inclination (T9-SPi), T1 spino-pelvic inclination (T1-SPi), T1 pelvic angle (TPA), spine-sacral angle (SSPA), and sagittal-vertical-axis angle (SVA-A) (the angle of the perpendicular line from the posterior edge of the S1 upper endplate to the center point of the C7 vertebra, with the right angle considered positive, thus converting the SVA distance value into an angle value to eliminate height differences) were measured.

### 3. Subject grouping

Based on pelvic incidence (PI): Group A <40°; Group B 40°≦PI<50°; Group C ≧50°. Based on the maximal thoracolumbar vertebral tilt(TLmax) (middle): Group A <106.5°; Group B 106.5°≦TLmax<114.5°; Group C ≧114.5°. The mean, standard deviation, and extreme values of each parameter were calculated, and chi-square tests were performed to compare and analyze the groups within the two classification types.

## Results

The measured and statistical parameters are presented in **Tables 1, 2, 3, and 4**.

**Table 1.**
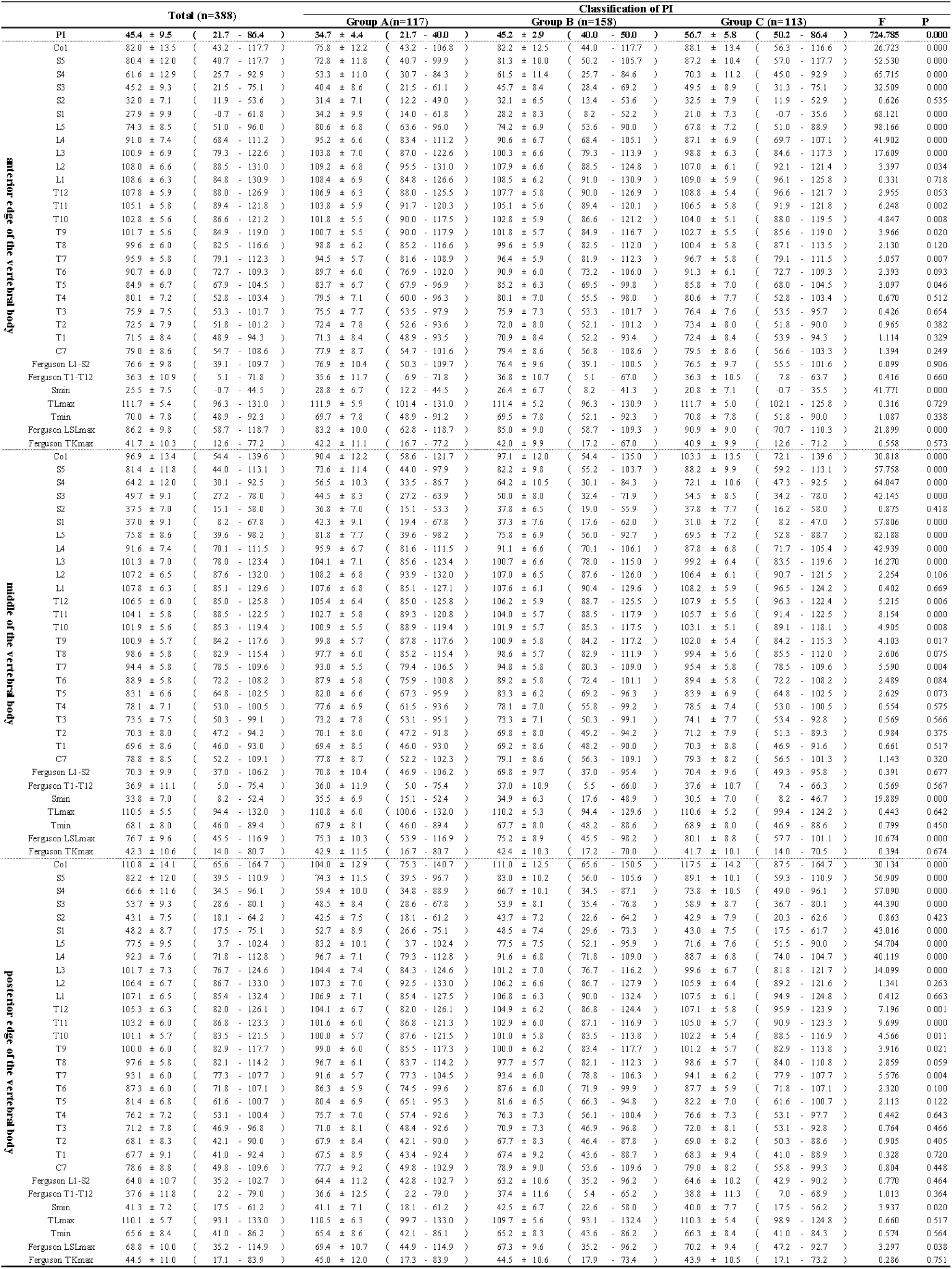
Classification based on PI - vertebral tilt and Ferguson angle of sagittal spinal aligment.

**Table 2.**
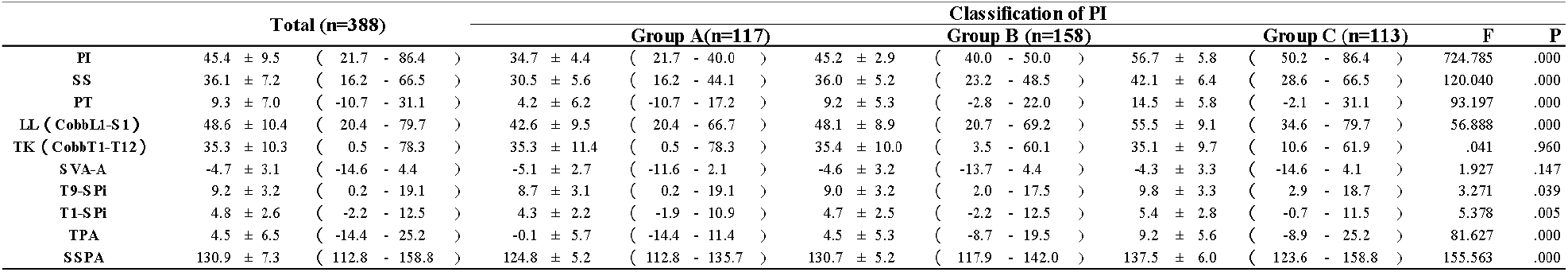
Classification based on PI - common sagittal spinopelvic parameters.

**Table 3.**
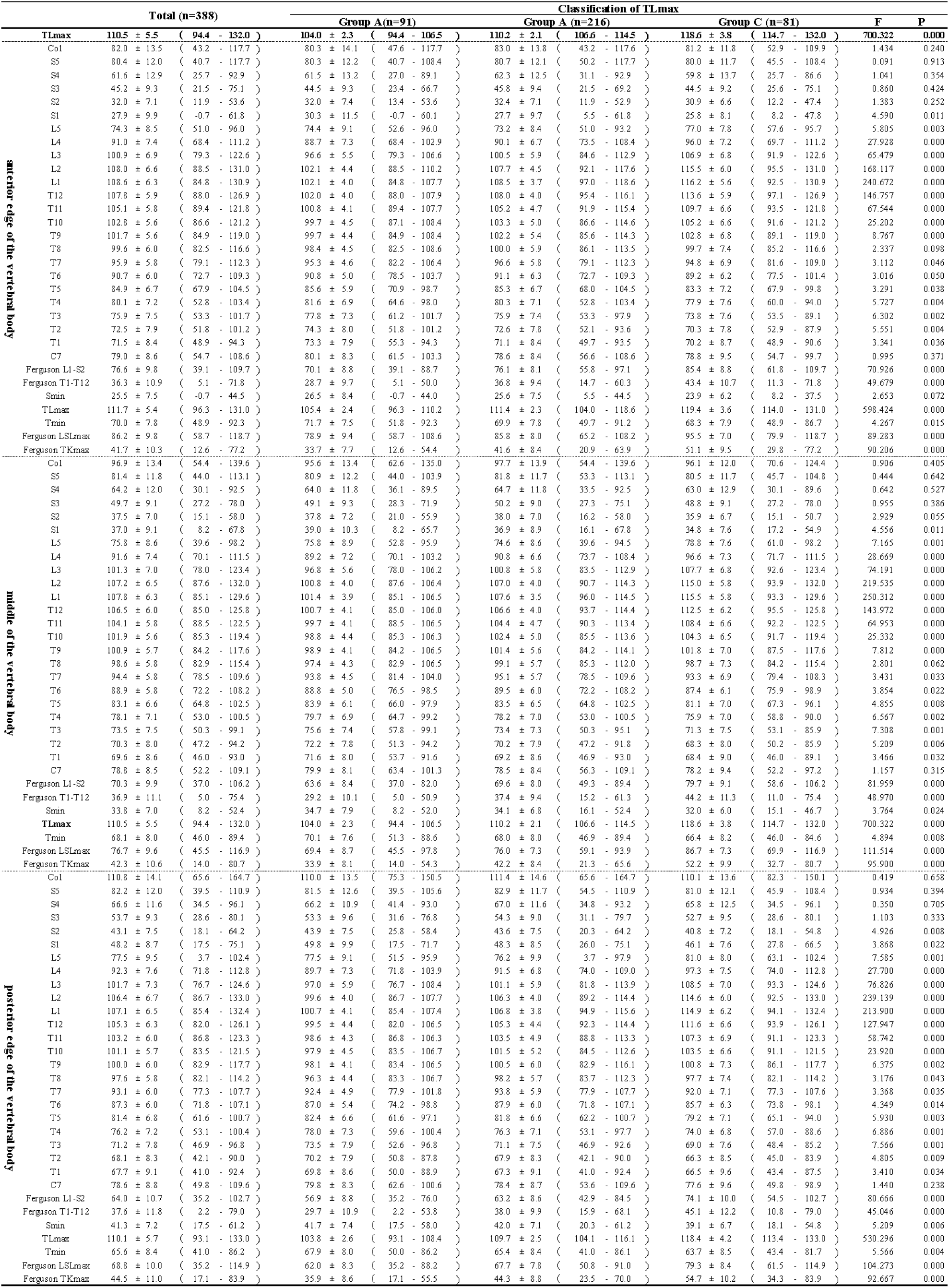
Classification based on TLmax - vertebral tilt and Ferguson angle of sagittal spinal aligment.

**Table 4.**
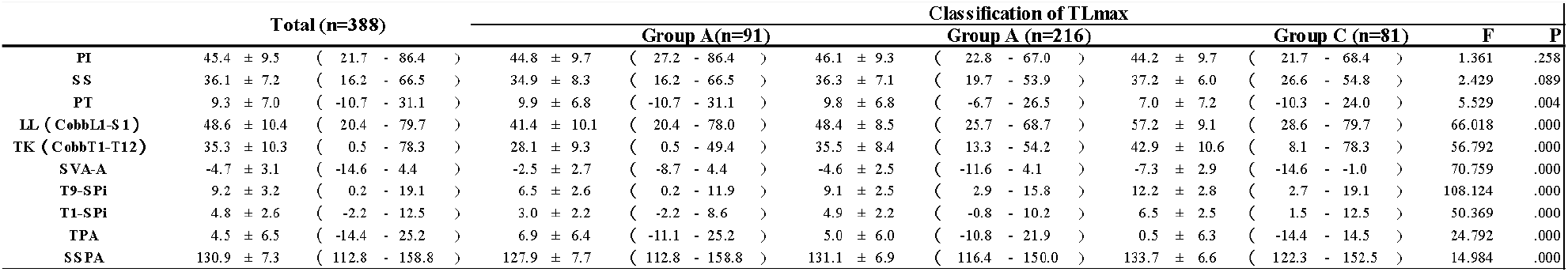
Classification based on TLmax - common sagittal spinopelvic parameters.

Based on PI grouping: The overall mean PI for all subjects was 45.4±9.5° (range 21.7-86.4°). Group A consisted of 117 subjects with a mean PI of 34.7±4.4° (range 21.7-40.0°); Group B had 158 subjects with a mean PI of 45.2±2.9° (range 40.0-50.0°); Group C included 113 subjects with a mean PI of 56.7±5.8° (range 50.2-86.4°). There were no significant statistical differences in the tilt of S2, L1, T8, T6, T5, T4, T3, T2, T1, C7, TLmax, Tmin among Groups A, B, and C; no significant statistical differences were found for Ferguson L1-S2, Ferguson T1-T12, FergusonTKmax; no significant statistical differences for Cobb T1-T12, SVA-A; however, other parameters showed statistical differences. **(Tables 1, 2)**

Based on TLmax grouping: The overall mean TLmax for all subjects was 110.5±5.5° (range 94.4-132.0°). Group A had 91 subjects with a mean TLmax of 104.0±2.3° (range 94.4-106.5°); Group B comprised 216 subjects with a mean TLmax of 110.2±2.1° (range 106.6-114.5°); Group C included 81 subjects with a mean TLmax of 118.6±3.8° (range 114.7-132.0°). There were no significant statistical differences in the tilt of Co1, S5, S4, S3, C7 among Groups A, B, and C, nor in PI, SS; a tendency for statistical differences was observed for S2, T8; other parameters showed statistical differences. **(Tables 3, 4)**

## Discussion

The evolution of humans from quadrupedal to bipedal locomotion, with the emergence of the “S” curve of the spine, which provided the necessary support for long-term upright activities with lower energy expenditure, facilitating their survival, reproduction, and development[3-6]. The emergence of the anterior convexity of the “S” curve is particularly important. It is commonly believed that the lordotic sequence is composed of the lumbar vertebraes and intervertebral discs, hence, most scholars report the angle between the upper endplates of L1 and S1 as the measurement index for this curve (lumbar lordosis, LL) [7]. However, from the perspective of spinal sequence, even in the normal population, the lordotic region does not strictly adhere to anatomical features, which has led to much confusion and debate in the assessment and application of the sagittal spinal aligment. Therefore, many scholars have redefined the “S” curve based on functional characteristics, no longer rigidly using the upper endplate of L1 as the boundary between the two bends, but instead, defining the boundary based on the inflection point of the spinal curvature. The definition of the inflection point is similar to the concept of the end vertebra commonly used in scoliosis, and its position does not depend on anatomical features. Consequently, the cephalic end of the lumbar lordosis may be located either proximally or distally on L1. Unlike the uncertainty of the cephalic end, the upper endplate of S1 is consistently used as the starting point of the lordosis, which may be related to the unique anatomical features of the sacrum.

However, contrary to previous studies that considered the sacrum as a whole kyphotic segment and S2 as the apex of kyphosis [12-14], Song et al.’s radiographic study demonstrated that, in terms of spinal sequence, the sacrum is not entirely kyphotic but is divided into a lordotic segment (S1-S2) and a kyphotic segment (S2-S5, Co1), with S1 and S2 effectively continuing the LL, and S2 should be considered the caudal end vertebrae of the lordosis [15]. Song et al. argued that, anatomically, S1 and S2 are within the tension band of the lumbar lordosis, as the erector spinae muscles terminates at the lamina and spinous process of S1 and S2, while the sacrotuberous ligament, sacrospinous ligament, and anococcygeal ligament insert at S3, S4, S5, and Co1, and their reverse tension determines the kyphotic characteristics of this region. Moreover, S2 is the stress core and rotational center of the sacroiliac joint, which is more consistent with the mechanical characteristics of an end vertebra rather than an apex vertebra [15]. Therefore, based on the radiographic study results and logical deductions from anatomy and biomechanics, S2 should be considered the caudal vertebra of the lordosis in the sagittal spinal aligment. The previous definition of the “S” curve’s lordotic range was incomplete, as it lacked a part of the sacral lordosis at its distal end. Thus, the definition of lumbosacral lordosis (LSL) seems more reasonable than that of lumbar lordosis (LL). The main reason for this omission is likely due to the limitations of the Cobb method in measuring the sacral region. For the lumbar spine, the Cobb method is relatively applicable due to the distinct boundaries provided by the larger intervertebral discs. However, for the sacral region, which undergoes fusion during development and significant morphological changes, making the Cobb method less applicable. This has led to the incompleteness of the radiographic evaluation for lordosis. To further explore the sagittal spinal aligment in humans, this study introduces the measurement using vertebral tilt and the Ferguson method, with a focus on studying LSL rather than LL.

The study compares two classifications of the sagittal spinal aligment in a healthy population: 1) classification based on pelvic incidence (PI); 2) classification based on the maximal thoracolumbar vertebral tilt(TLmax).

### 1. Classification based on Pelvic Incidence (PI)

According to the anatomical segmentation, the study results show that for groups with different types of PI, there are no statistically significant differences in the vertebral tilt of the transitional vertebrae S2, L1, T1 for the “S” curve anatomical segmentation (Groups A, B, C), nor are there differences in the lumbosacral lordosis (LSL) Ferguson L1-S2 and thoracic kyphosis (TK) Ferguson T1-T12. These results imply that PI does not significantly affect the overall magnitude of the lumbosacral lordosis and thoracic kyphosis; in other words, the overall curvature of the “S” curve of the spine is essentially unaffected by PI according to anatomical segmentation. **(Table 1, Figure 1)**

**Figure 1.**
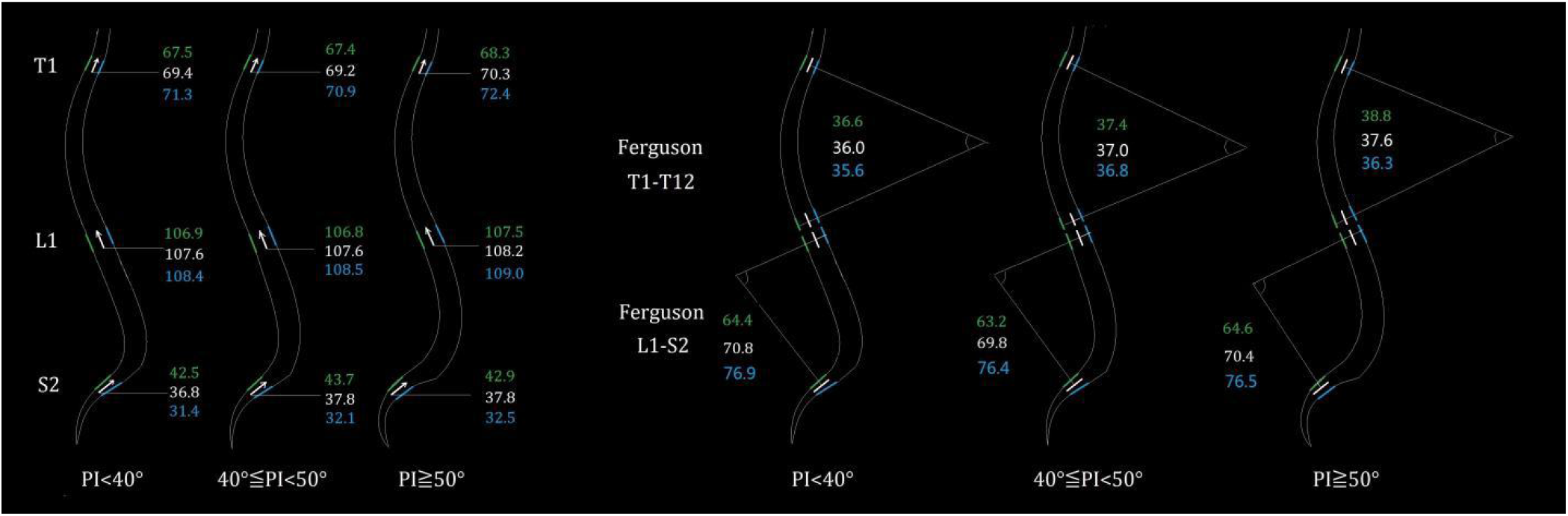
Classification based on PI **-** anatomical segmentation

Based on functional segmentation, there are no significant statistical differences in TLmax and Tmin; Ferguson TKmax also shows no significant statistical difference. There is a statistical difference in the Smin and Ferguson LSLmax. The study results indicate that it is more common for the anterior edge of Smin to be located at S1, while the posterior edge of Smin is mainly concentrated at S2, with the middle part of the vertebrae falling between the two. Unlike the thoracolumbar vertebrae, which are rectangular in shape, the sacral vertebrae are trapezoidal, with S1 having a greater trapezoidal gradient than S2. The variation in the anterior edge tilt is greater than that of the posterior edge, which could significantly affect the difference between measurement results and the true sequence. As the results show, the anterior edge of Smin has a greater variation than the middle and posterior edges, and although there is a statistical difference in the posterior edge (0.01<P<0.05), the significance is lower than that of the anterior and middle edges (P<0.001), and the mean difference is small, even within the acceptable measurement error range. Based on these inferences, the author believes that due to the specific morphology of S1, even with the use of vertebral tilt measurements and the Ferguson method, the measurement results may not accurately reflect its sagittal aligment, and this could also likely lead to the statistical differences in Smin and Ferguson LSLmax observed in this study. Although not proven, based on the logical analysis of the results, the author leans towards the belief that the true Smin and Ferguson LSLmax also do not show statistical differences. If that were the case, according to functional segmentation, the overall curvature of the spinal “S” curve is also unaffected by PI. **(Table 1, Figure 2)**

**Figure 2.**
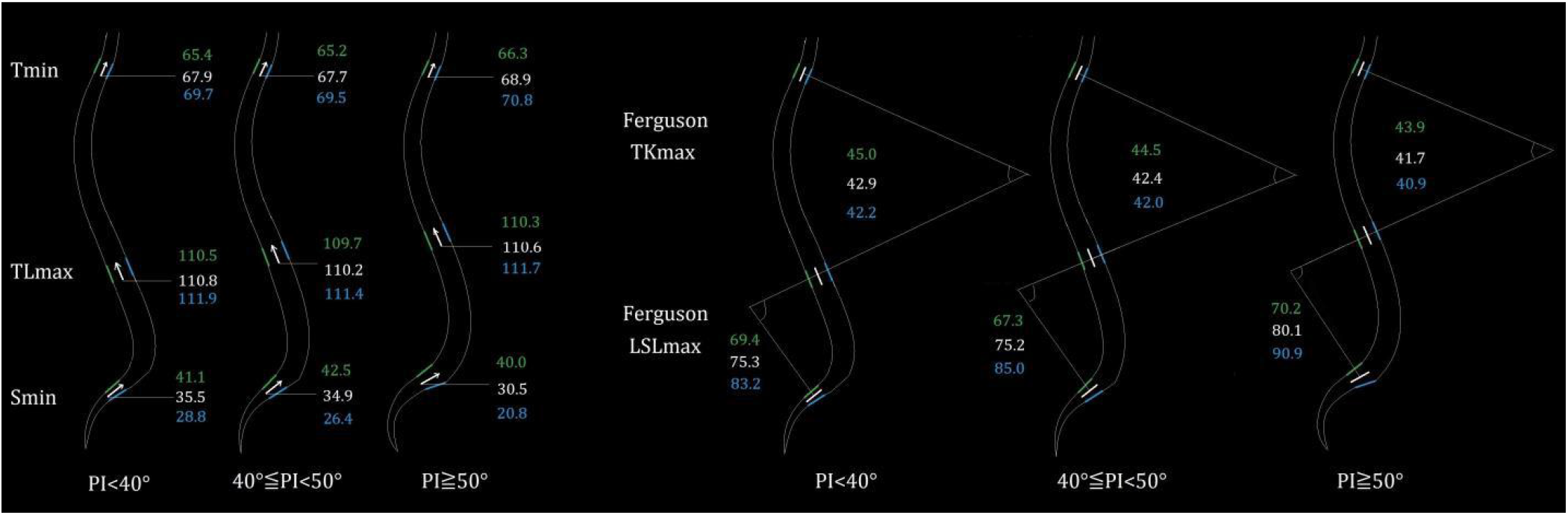
Classification based on PI - functional segmentation

The study results also show that for groups with different types of PI, there are varying degrees of statistical differences or trends in S1, L5, L4, L3, L2, T12, T11, T10, T9, T7, with more significant differences in the lumbosacral segment. This means that although the overall curvature of the “S” curve of the spine is the same across different PI groups, there are differences in local curvature. The author will discuss the mechanisms and logic behind this in subsequent articles.

### 2. Classification based on the maximal thoracolumbar vertebral tilt (TLmax)

Regardless of whether anatomical or functional segmentation is considered, this study shows that for groups with different TLmax, there are significant statistical differences in the tilt of the transitional vertebrae S2, L1, T1 for the “S” curve anatomical segmentation, as well as the transitional vertebrae Smin, TLmax, Tmin for functional segmentation (Groups A, B, C). There are also significant statistical differences in LSL, (Ferguson L1-S2), TK (Ferguson T1-T12), Ferguson LSLmax, and Ferguson TKmax. These results imply that using TLmax as a classification reference can effectively differentiate the overall curvature of the “S” curve of the human spine.**(Table 3, Figure 3,4)**

**Figure 3.**
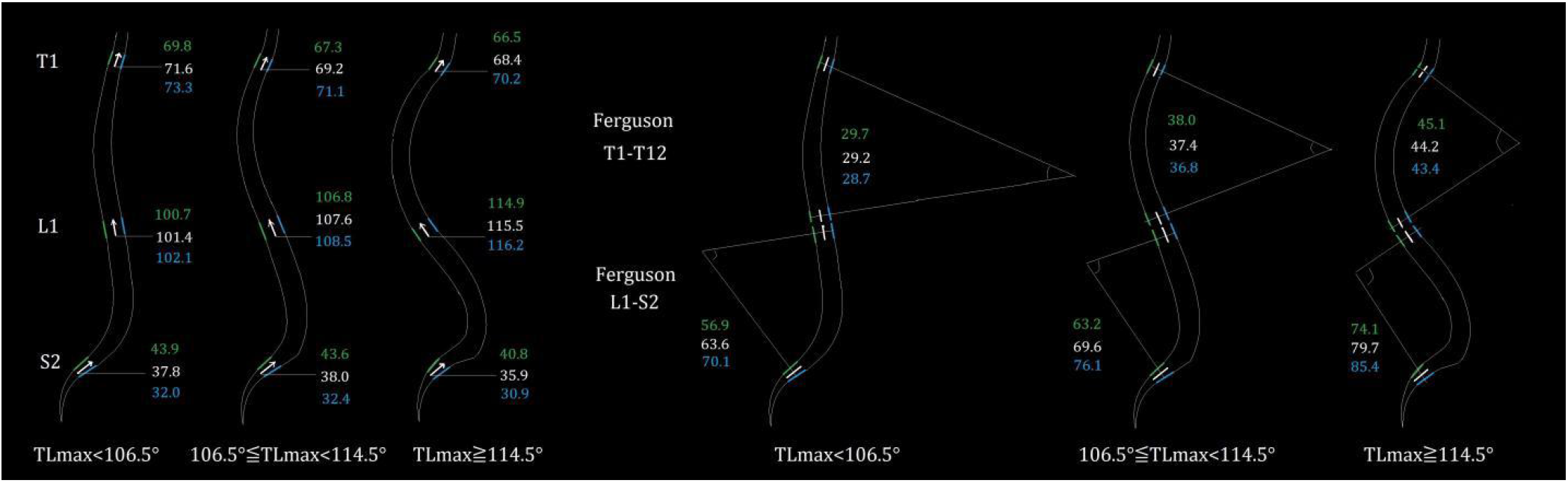
Classification based on TLmax - functional segmentation

**Figure 4.**
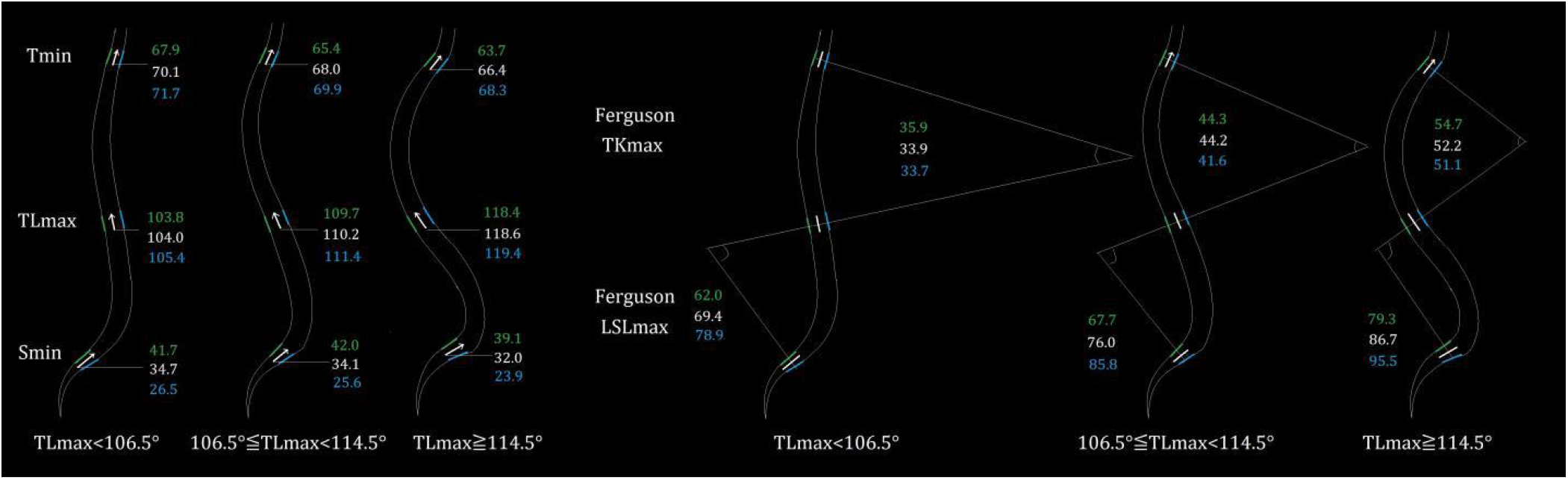
Classification based on TLmax - anatomical segmentation

At the same time, S3 and C7, as adjacent segments of the “S” curve, show no significant statistical differences in their tilts. This result means that the curvature magnitude of the “S” curve does not have a significant impact on the regional range of curve. Additionally, there are no significant statistical differences in PI and SS across the groups of this classification method, which further proves that PI, and even SS, do not substantially decide the overall curvature magnitude of the spinal “S” curve.

The classification method using TLmax as a reference provides an alternative perspective for our research on the sagittal balance of the human spine, which could potentially influence the diagnosis and treatment strategies for disorders of the spinal sagittal aligment.

#### Limitations of the study

1. The study utilized a multicenter sample, and for a subset of participants, data on gender and precise age were absent. 2. PI is a morphological parameter. The author had intended to contrast and categorize using newly developed morphological parameters, but the description and interpretation of the parameter would necessitate a substantial exposition. To ensure a straightforward and coherent narrative, this paper provisionally employs the positional parameter TLmax as an interim classification method to avoid reader confusion. 3. The sample sizes for the two categorical groups in this study were not uniform. This is because uniform sample sizes across all categories and groups did not yield sufficiently significant statistical outcomes, which could potentially perplex readers. The study’s approach to sample categorization aims to clarify results but may inherently introduce a degree of logical stringency imperfection.

## Conclusion

Pelvic Incidence (PI) does not influence the overall magnitude of lumbosacral lordosis(LSL) and thoracic kyphosis(TK). The sagittal spinal classification based on PI reveals no difference in the overall curvature of the spinal “S” shape. The maximum thoracic kyphosis (TLmax) significantly affects LSL and TK, and it can be considered a straightforward radiological parameter for distinguishing the overall curvature of the spinal “S” shape.

## Data Availability

All data produced in the present study are available upon reasonable request to the authors

